# City-level SARS-CoV-2 sewage surveillance

**DOI:** 10.1101/2020.10.19.20215244

**Authors:** Karin Yaniv, Marilou Shagan, Esti Kramarsky-Winter, Merav Weil, Victoria Indenbaum, Michal Elul, Oran Erster, Alin Sela Brown, Ella Mendelson, Batya Mannasse, Rachel Shirazi, Satish Lakkakula, Oren Miron, Ehud Rinott, Ricardo Gilead Baibich, Iris Bigler, Matan Malul, Rotem Rishti, Asher Brenner, Yair E. Lewis, Eran Friedler, Yael Gilboa, Sara Sabach, Yuval Alfiya, Uta Cheruti, Nadav Davidovitch, Natalya Bilenko, Jacob Moran-Gilad, Yakir Berchenko, Itay Bar-Or, Ariel Kushmaro

**Affiliations:** Avram and Stella Goldstein-Goren, Department of Biotechnology Engineering, Ben-Gurion University of the Negev, Beer-Sheva, Israel; Central Virology Lab, Ministry of Health, Sheba Medical Center, Israel; The Ilse Katz Center for Meso and Nanoscale Science and Technology, Ben-Gurion University of the Negev, Be’er Sheva 8410501, Israel; Department of Industrial Engineering and Management, Ben-Gurion University of the Negev, Beer-Sheva 84105, Israel; Department of Health Systems Management, School of Public Health, Faculty of Health Science, Ben-Gurion University of the Negev, Beer-Sheva, Israel; Faculty of Medicine, Technion-Israel Institute of Technology, Israel; Faculty of Civ. and Env. Eng., Technion-Israel Inst. of Technology; Haifa 32000, Israel; School of Public Health, Sackler Faculty of Medicine, Tel-Aviv University, Tel-Aviv, Israel; KANDO, Environment Services Ltd, Tsor St 8, Kokhav Ya’ir Tzur Yigal, Israel; Unit of Environmental Engineering, Ben-Gurion University of the Negev, Beer-Sheva, Israel

## Abstract

The COVID-19 pandemic created a global crisis impacting not only healthcare systems, but also world economies and society. Recent data have indicated that fecal shedding of SARS-CoV-2 is common, and that viral RNA can be detected in wastewater. This suggests that wastewater monitoring is a potentially efficient tool for both epidemiological surveillance, and early warning for SARS-CoV-2 circulation at the population level. In this study we sampled an urban wastewater infrastructure in the city of Ashkelon, Israel, during the end of the first COVID-19 wave in May 2020 when the number of infections seemed to be waning. We were able to show varying presence of SARS-CoV-2 RNA in wastewater from several locations in the city during two sampling periods. This was expressed as a new index, Normalized Viral Load (NVL), which can be used in different area scales to define levels of virus activity such as red (high) or green (no), and to follow morbidity in the population at tested area. Our index showed the rise in viral load between the two sampling periods (one week apart) and indicated an increase in morbidity that was evident a month later in the population. Thus, this methodology may provide an early indication for SARS-CoV-2 infection outbreak in a population before an outbreak is clinically apparent.

**HIGHLIGHTS:**

- Detecting the presence of SARS-CoV-2 virus RNA in urban wastewater
- The city sewer system may provide an early indication for SARS-CoV-2 infection and may be used as early warning for SARS-CoV-2 outbreaks
- NVL index defines various infected urban zones from red (high) to green (low)

**Graphical abstract:** 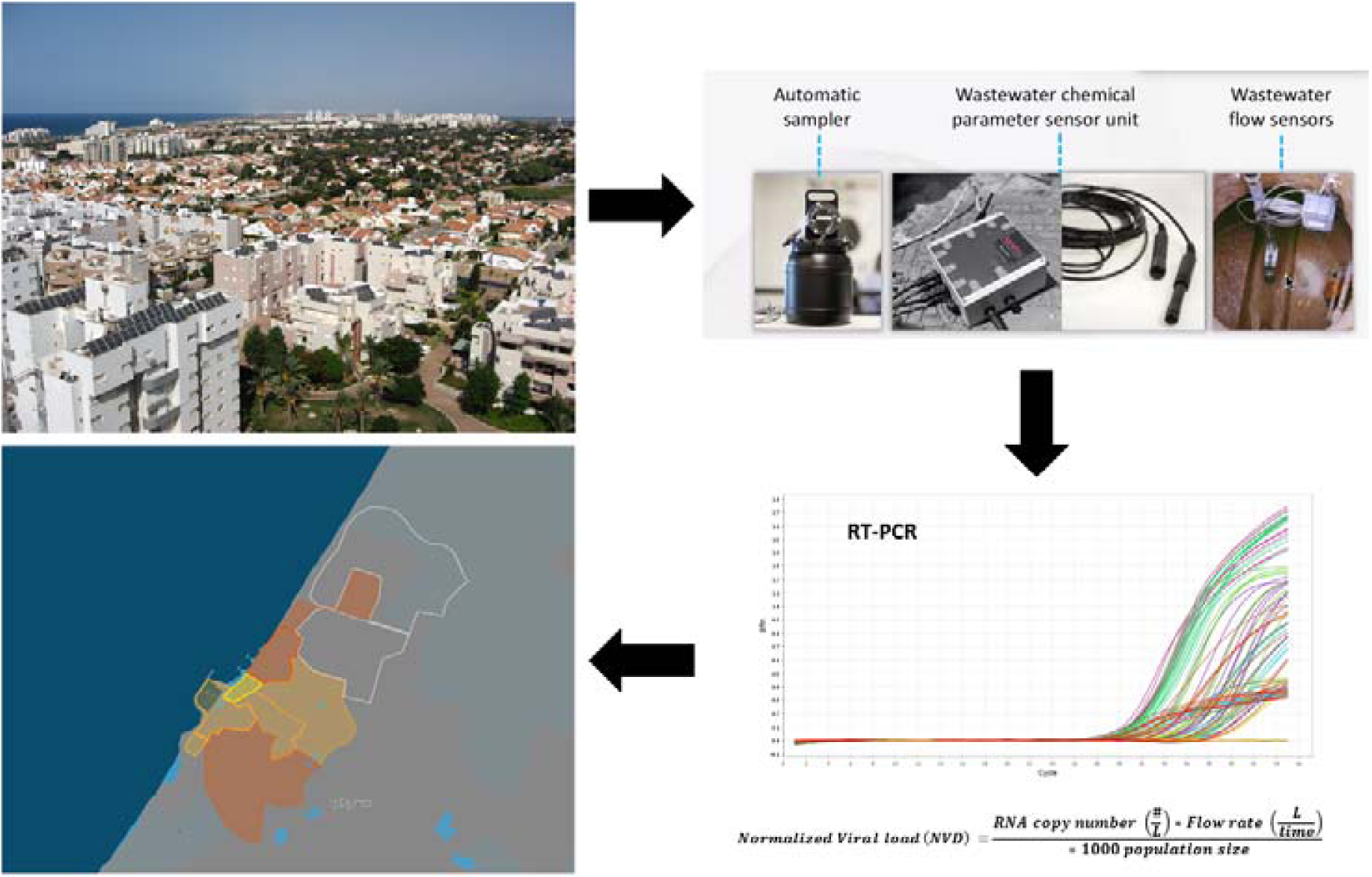

## Introduction

Monitoring of urban wastewater systems can provide an efficient tool for infectious disease surveillance and can be used as an early warning system. This strategy has been used for the past decades for monitoring infectious diseases such as Polio^1^, antibiotic resistance^2^ and other microbial pathogens^3^. SARS-CoV-2 emerged during late 2019 and has resulted in an ongoing pandemic that to date is difficult to eradicate. As vaccines are not going to be widely available soon, it is important to find novel methods for monitoring disease activity. Recent reports have shown that SARS-CoV-2 is shed through human stool^5,6^ and can be detected in wastewater, as evident from studies worldwide^7^, including Israel^8^. Therefore, environmental surveillance of SARS-CoV-2 in sewage may provide a reliable monitoring tool for assessing disease activity and resurgence. The reliable assessment of the viral load of a population using this methodology is dependent upon many variables including the city’s sewer system topology, population, and usage. This study aims to develop an urban-sewer sampling strategy for SARS-CoV-2 monitoring in different populated areas enabling geographical localization of COVID-19 outbreaks.

## Materials and methods

### Sampling

The study was performed in the city of Ashkelon, a Southern coastal city in Israel having a population of approximately 150,000 inhabitants. Ten continuous sewage-sampling units were installed in Ashkelon’s wastewater system at nine selected sewer manholes and one at the inlet of the wastewater treatment plant (WWTP). The automated composite samplers were activated during two sampling campaigns, the first during 17-19 of May 2020 and the second on the 25^th^ of May 2020. Sampling was carried out for 8 hours, from 6:00-14:00 or 07:00-15:00 at each point. The chosen sampling areas included several neighborhoods along the coast of Ashkelon (tagged as 1 to 4 in Figure 1). The sampling points included only household wastewater. They were connected to a central sewer line that was also sampled in designated sewer junction manholes (A to E in Figure 1). This central sewer line eventually drains into the municipal WWTP (designated as point H) that treats the city wastewater. For 1 to 4 neighborhoods, the estimated population sizes were 13,500; 5,600; 2,500; 12,500 respectively, based national census data (the Central Bureau of Statistic, https://www.gov.il/en/departments/central_bureau_of_statistics).

**Figure 1.**
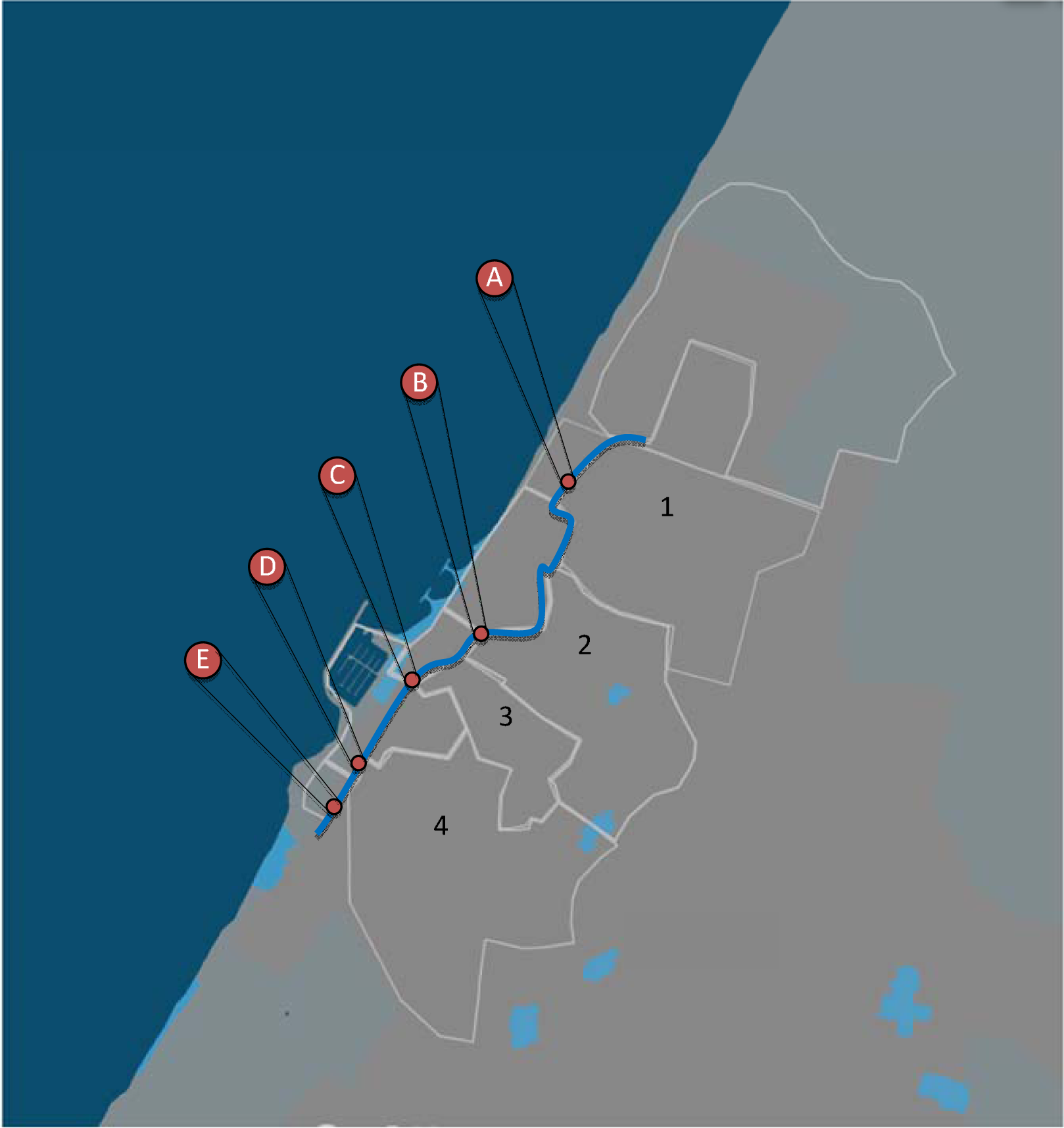
Deployment of sampling units in the city of Ashkelon.

### Sample RNA extraction and analysis

Raw sewage samples (1L composite sample) were immediately transferred to the lab under chilled conditions. The samples were kept at – 20°C until processed. Prior to subsampling, the bottles were shaken vigorously and left to rest for 10 minutes. Subsamples of 0.2 mL raw sewage (in duplicate) were then transferred directly into NucleoSpin RNA lysis buffer for RNA extraction (Macherey Nagel, Germany). For creating a standard curve, we used a plasmid containing the full SARS-CoV-2 N gene sequence isolated from Wuhan-Hu-1 (GenBank: NC_045512.2). We prepared standard curves of plasmid log copy number verses Ct value using serial dilutions of the plasmid. We then performed linear regression between the log copy number and the Ct values from the RT-qPCR results. Using the linear equation, we calculated copy number of N1 gene in sewage samples reported in this study. RT-qPCR amplification was performed according to user manual recommendation using CDC’s N1 and N2 primers and probe sets and One Step PrimeScript III RT-qPCR mix (TAKARA, Japan). RT-qPCR amplification was executed using Step One Plus real-time PCR system (Applied Biosystems, Thermo Scientific). In parallel to the N1 test, each RNA sample was spiked with N gene plasmid in known concentrations to rule out any amplification inhibition. A second quality control step was carried out by adding MS2 phage to the lysis buffer step as process control for RNA extraction. The analytical LOD (limit of detection) for N gene target was determined to be < 10 target copies per qPCR reaction.

### Viral load calculation

Viral copy number per 1L was calculated using the N1 standard curve equation (Figure S1). If Ct value for N1 was ND, then N2 Ct value was used for the calculation. A new index, namely Normalized Viral Load (NVL) was defined. It is expressing the estimated number of SARS-CoV-2 RNA copies per 1,000 people. It is based on quantification of the cumulative number of copies during several hours of composite wastewater sample collection (8 hours in this study) in a specific location, within a steady timeframe of a day (see equation1). It can be fitted to various scales such as a street, a quarter, a small town, or a large city. Sampling location, duration, and method (preferably composite) should be unified to enable reliable use of the NVL. If the sampling is extended to 24 hours, then the NVL can be expressed as number of SARS-CoV-2 RNA copies per 1,000 people per day. Nevertheless, even sampling of several hours during a steady timeframe can serve as a reliable relative index to be assessed chronologically. This index can assist define levels of virus activity in a semi-quantitative fashion (e.g. using a ‘traffic-light’ scheme), and to follow morbidity of the population in the tested area.

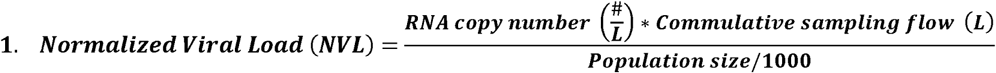

## Results and discussion

Ashkelon was chosen for surveillance of SARS-CoV-2 in sewage due to a relatively low COVID-19 prevalence during the time of study following a national lockdown period during April 2020 so that by beginning of May life gradually returned to normal. Ashkelon is a metropolitan area with approximately 150,000 residents. The urban sewage system design was composed from 10 continuous sewage-sampling units. During May 2020 we deployed continuous sampling devices in nine major sewer manholes in the city’s wastewater system and in the city wastewater treatment plant (WWTP). Sewage samples were tested for SARS-CoV-2 RNA and chemical analysis of these samples were performed as well including TOC, TN, and pH (see Table S1).

In this study we carried out two sampling campaigns during May 2020, a period with limited reports of COVID-19 in the city and with a very low prevalence of COVID-19 in all of Israel. This low prevalence was achieved due to the national lockdown period during April 2020 so that by beginning of May life returned gradually to normal. Despite the fact that no new COVID-19 positive cases had been reported in Ashkelon to the Ministry of Health during this time, we found traces of the SARS-CoV-2 RNA in sewage originating from the different sampling manholes in the city (Figure 2). Figure 2a presents the accumulation of active COVID-19 cases in Ashkelon as quantified by healthcare diagnostics and reported by the ministry of health. The arrows in the figure indicate the two sampling efforts during May 2020. During the first sampling campaign, two sampling locations (A and 1) were positive for SARS-CoV-2 with normalized viral load (NVL) of 3.63·10^12^ and 1.29·10^13^ (copy number/1000 person) respectively. The remaining sampling point were negative (Not Detected, ND) including point H which is the inlet of the WWTP. These results indicate the dilution effect in the sewer system and suggests a low sensitivity of the method when sampling is applied only to the WWTP or larger geographical areas, especially when the prevalence is low and/or highly localized.

**Figure 2.**
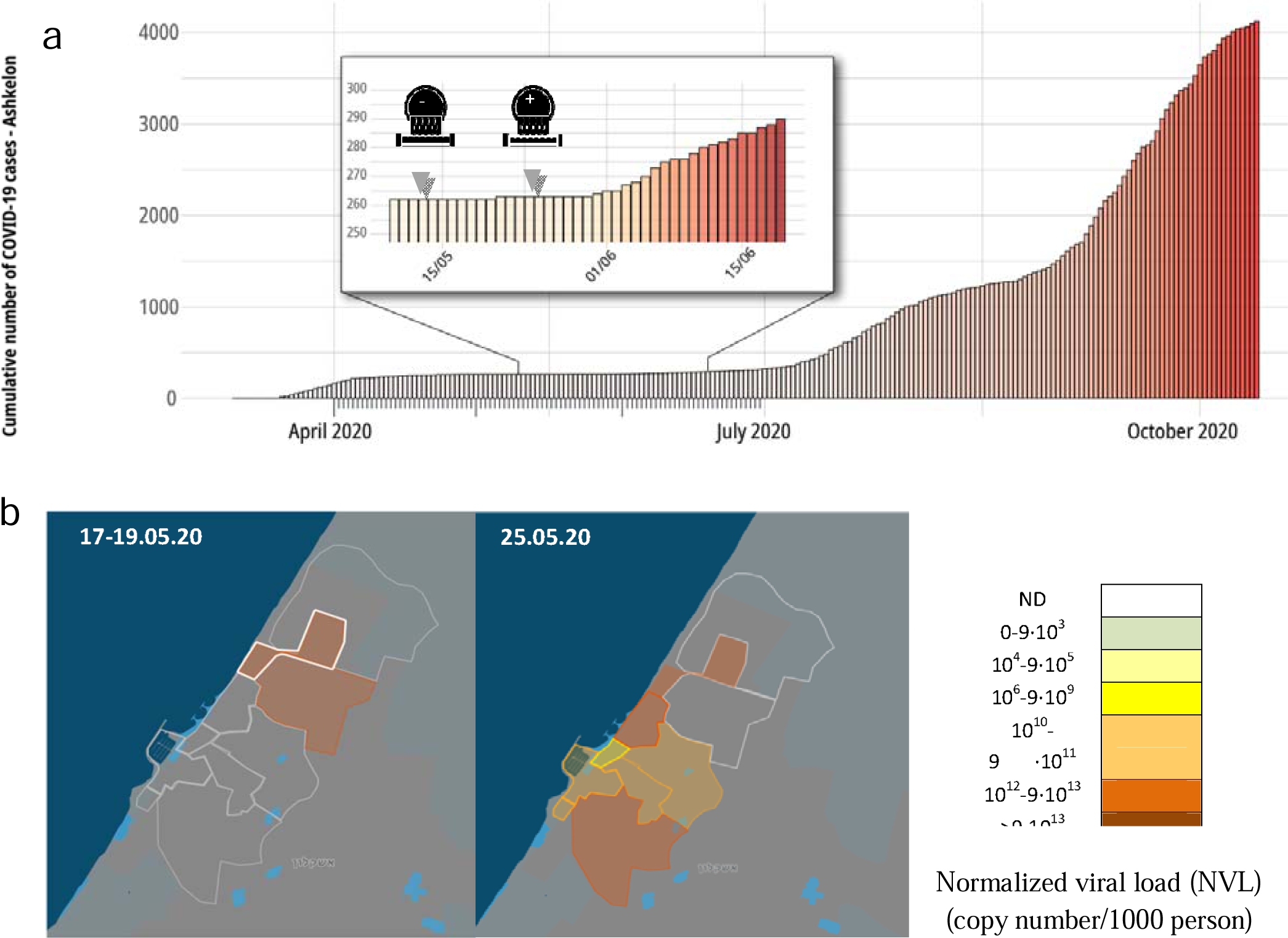
(a) Active COVID-19 cases in Ashkelon during March until August 2020. Black dash line presents the beginning of the second COVID-19 outbreak (reported by health care diagnostics). Arrows in the zoomed-in box, represent the sampling campaigned dates in May 2020 and the illustration icons represent the SARA-CoV-2 detection results at the WWTP. (b) Normalized SARS-CoV-2 viral loads (NVL) calculated from wastewater collection campaign from different geographic locations at the city of Ashkelon.

In the second sampling campaign all sampling points were positive, with NVL between 5.9·10^11^ to 2.77·10^13^ (copy number/1000 person). Unfortunately, due to technical problem point 1 sample is missing from the second campaign. Interestingly, the viral load in point A increased by an order of magnitude from NVL of 3.63·10^12^ to 2.77·10^13^ (copy number/1000 person) between the two sampling campaigns. Figure 2b demonstrates, by color code, the NVL in each of the sampling manholes, recording the profound changes in viral load during the two sampling campaigns. While two time points are not enough to establish a trend, it is important to point out that we detected an abnormal signal about one month prior to the July COVID-19 outbreak of the second wave of in the city (seen in figure 2a). When focusing on the May sampling (Figure 2a), one can observe a relationship to reported COVID-19 cases. The low morbidity in the city reported during May, particularly at the end of the month, might have been an under estimation of COVID-19 cases due to limited testing efforts and due to asymptomatic cases (and therefore untested per testing guidelines at the time of study). The presence of the virus in the sewage despite the low prevalence reported in May attests to the sensitivity of the method. Indeed, a resurgence in COVID-19 cases became evident in the city’s population in June-July 2020.

Understanding the dynamics of SARS-CoV-2 in human excreta could lead to efficient monitoring and surveillance of this virus as well as to underpin the important of such environmental surveillance^9^. Our results also demonstrate the need for an inner-city sewer sampling design to overcome dilution effect that can eventually affect the sensitivity and validity of this surveillance method. Such an inner-city sewer study may assist in geographical localization of disease hotspots. Multiple sampling efforts within a city could allow assessment of virus spread in each area and assess the proportion of areas with virus activity in a city, thus predicting imminent resurgences.

In conclusion, we present a proof-of-concept study demonstrating the feasibility of SARS-CoV-2 RNA detection in raw sewage originating from the city sewer system that reflects virus circulation in the assessed area. This approach should be further studied and validated as an early warning for SARS-CoV-2 resurgence in urban settings and as a national decision support tool^10,11^.

## Data Availability

All data is presented in the manuscript

## Author Contributions

K.Y executed all SARS-CoV-2 molecular analyses, calculations and wrote this manuscript. M.S and S.L extracted RNA samples. E. K.W performed language editing. M.W, V.I, M.E, O.E, A.S.B, E.M, B.M and R.S contributed to method development. O.M, E.R, R.G.B, I.B, M.M, R.R, Y.E.L, N.D, J.M.G and Y.B were in charge of sampling design and execution. J.M.G, E.F and N.D provided critical comments for the manuscript. A.B participated in calculations and data analysis. E.F, Y.G, S.S, Y.A and U.C performed wastewater chemical analyses. I.B.O and A.K wrote this manuscript and took part in experimental design.

## Abbreviations

NVL: (Normalized Virus Load)
WWTP: (Wastewater Treatment Plant)
CDC: (the Center of Disease Control)
RT-qPCR: (Real Time quantification Polymerase Chain Reaction)
LOD: (Limit of Detection)
ND: (Not Detected)

## Funding Sources

We would like to acknowledge funding from Ben Gurion University, The Corona Challenge Covid-19 (https://in.bgu.ac.il/en/corona-challenge/Pages/default.aspx), The Milner Foundation for Corona Study (Technion) and funding from the Israeli ministry of Health.

## Displayed equations

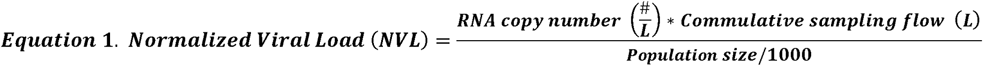

## Supporting Information

### Wastewater analysis

Raw sewage (1L composite sample) was irradiated for two hours using LP-UV lamp (7 mW/cm2, VL_230.G, Lamp part number T-30.C, Cole Parmer Instrument Co., USA.). During the irradiation process, samples were continuously mixed in order to assure that the samples are properly inactivated. Samples were filtered through 90 mm GF/A glass fiber filter (Whatman, UK). Filtered samples were analyzed for total organic carbon (TOC) and total nitrogen (TN), using total organic carbon analyzer (TOC-V CPH, equipped with total nitrogen measuring unit, TNM-1; Shimadzu, Japan).

**Table 1:** Results for SARS-CoV-2 virus concentration in different sampling point in Ashkelon city and TN, flow rate, population values.

| Sample name | TOC (mg/L) | TN (mg/L) | Flow (m<sup>3</sup>/hr) | Cummulative sampling flow (m<sup>3</sup>) | Sample date | NCOV-N1 | NCOV-N2 | NCOV-N4 | MS2 | Spike N gene block (N1 OR N2) | N1_Virus concentration per L | N2_Virus concentration per L | Population | Viral load_Copy number per day per person | Viral load_Copy number per day per 1000person |
| --- | --- | --- | --- | --- | --- | --- | --- | --- | --- | --- | --- | --- | --- | --- | --- |
| H | 128 | 92 | 11931.14 | 95449.09 | 25.05 | 35.22 | ND | 36.04 | 29.91 | 26.53 | 9.02E+06 |  | 1.50E+05 | 5.74E+09 | 5.74E+12 |
| E | 131 | 99 | 4725.14 | 37801.08 | 25.05 | ND | 34.66 | 35.32 | 29.35 | 26.56 |  | 2.67E+06 | 4.85E+04 | 2.08E+09 | 2.08E+12 |
| D | 119 | 94 | 3981.17 | 31849.33 | 25.05 | 35.89 | ND | 35.57 | 29.89 | 26.55 | 5.67E+06 |  | 3.58E+04 | 5.04E+09 | 5.04E+12 |
| C | 145 | 103 | 3127.51 | 25020.05 | 25.05 | ND | 36.4 | 39.03 | 29.99 |  |  | 7.73E+05 | 3.28E+04 | 5.90E+08 | 5.90E+11 |
| B | 110 | 76 | 2581.91 | 20655.24 | 25.05 | 35.3 | 36.34 | ND | 29.90 | 26.76 | 8.53E+06 |  | 2.70E+04 | 6.53E+09 | 6.53E+12 |
| A | 124 | 101 | 1197.61 | 9580.85 | 25.05 | 33.18 | 36.41 | ND | 28.36 | 26.79 | 3.71E+07 |  | 1.28E+04 | 2.77E+10 | 2.77E+13 |
| 4 | 122 | 89 | 743.97 | 5951.75 | 25.05 | 34.86 | ND | ND | 29.34 | 26.67 | 1.16E+07 |  | 1.25E+04 | 5.51E+09 | 5.51E+12 |
| 3 | 118 | 84 | 853.66 | 6829.29 | 25.05 | 36.66 | 37.4 | ND | 29.09 | 26.26 | 3.32E+06 |  | 2.50E+03 | 9.08E+09 | 9.08E+12 |
| 2 | 194 | 123 | 545.60 | 4364.80 | 25.05 | 35.24 | ND | ND | 30.33 | 26.33 | 8.89E+06 |  | 5.60E+03 | 6.93E+09 | 6.93E+12 |
| H | 133 | 91 | 12207.49 | 97659.91 | 19.05 | ND | ND | ND | 28.63 | 26.41 |  |  | 1.50E+05 |  |  |
| E | 122 | 92 | 3884.51 | 31076.10 | 19.05 | ND | ND | ND | 30.12 | 26.74 |  |  | 4.85E+04 |  |  |
| D | 131 | 97 | 4060.91 | 32487.28 | 18.05 | ND | ND | ND | 30.33 | 25.51 |  |  | 3.58E+04 |  |  |
| C | 135 | 94 | 3210.75 | 25685.99 | 19.05 | ND | ND | ND | 29.48 | 25.61 |  |  | 3.28E+04 |  |  |
| B | 91 | 60 | 1799.97 | 14399.78 | 18.05 | ND | ND | ND | 28.79 | 26.21 |  |  | 2.70E+04 |  |  |
| A | 143 | 96 | 516.16 | 4129.27 | 17.05 | 34.9 | 33.8 | 35 | 30.10 | 25.33 | 1.13E+07 | 4.93E+06 | 1.28E+04 | 3.63E+09 | 3.63E+12 |
| 4 | 140 | 98 | -363.40 | -2907.21 | 18.05 | ND | ND | ND | 29.14 | 25.96 |  |  | 1.25E+04 |  |  |
| 3 | 108 | 78 | 868.96 | 6951.69 | 19.05 | ND | ND | ND | 30.20 | 27.01 |  |  | 2.50E+03 |  |  |
| 2 | 140 | 99 | 1341.05 | 10728.43 | 19.05 | ND | ND | ND | 29.51 | 26.43 |  |  | 5.60E+03 |  |  |
| 1 | 146 | 97 | 1283.81 | 10270.51 | 18.05 | 34.31 | 33.96 | ND | 28.94 | 27.05 | 1.69E+07 |  | 1.35E+04 | 1.29E+10 | 1.29E+13 |

**Figure S1:**
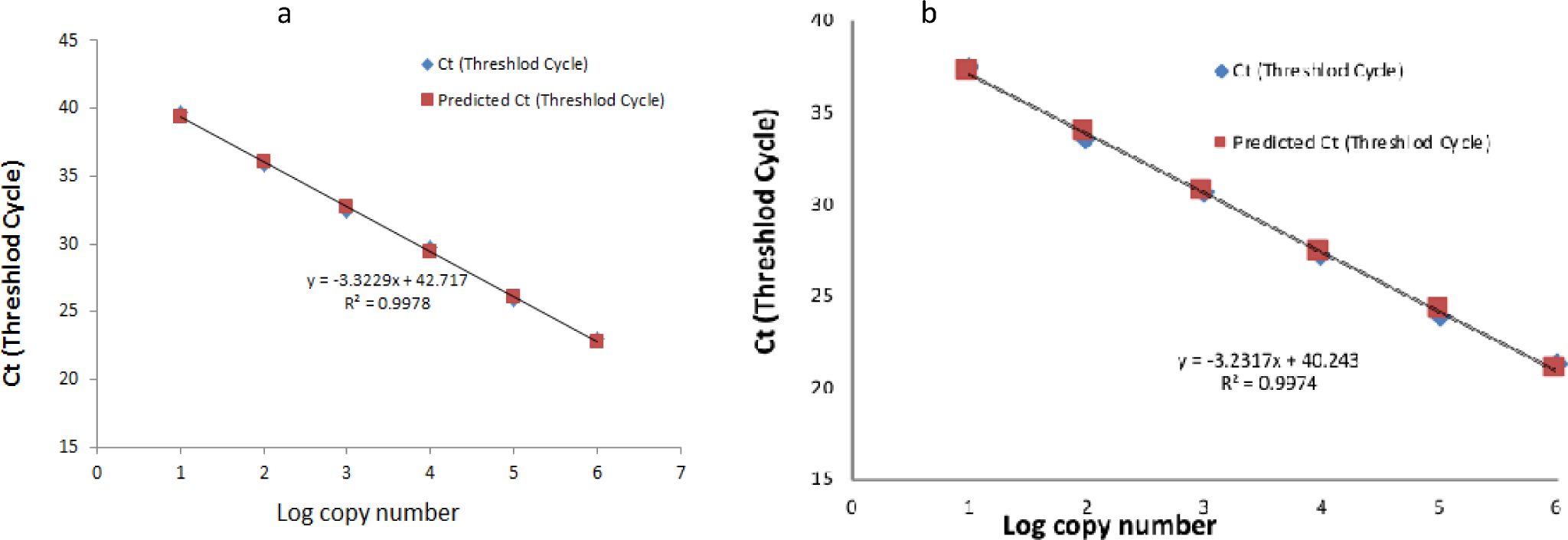
calibration curve for CDC’s N1 (a) and N2 (b) primers and probe set. Linear regression between the log copy number of SARA-CoV-2 N gene plasmid and the Ct values from the RT-PCR amplification.

## Notes

### Competing Interest Statement

The authors have declared no competing interest.

### Author Declarations

Ben Gurion University

## References

(1) Manor, Y., Shulman, L. M., Kaliner, E., Hindiyeh, M., Ram, D., Sofer, D., Moran-Gilad, J., Lev, B., Grotto, I., Gamzu, R., Mendelson, E. Intensified Environmental Surveillance Supporting the Response to Wild Poliovirus Type 1 Silent Circulation in Israel, 2013. Eurosurveillance 2014, 19 (7), 1–10. https://doi.org/10.2807/1560-7917.ES2014.19.7.20708.

(2) Hendriksen, R. S., Munk, P., Njage, P., van Bunnik, B., McNally, L., Lukjancenko, O., Röder, T., Nieuwenhuijse, D., Pedersen, S. K., Kjeldgaard, J., Kaas, R. S., Clausen, P. T. L. C., Vogt, J. K., Leekitcharoenphon, P., van de Schans, M. G. M., Zuidema, T., de Roda Husman, A. M., Rasmussen, S., Petersen, B., Amid, C., Cochrane, G., Sicheritz-Ponten, T., Schmitt, H., Alvarez, J. R. M., Aidara-Kane, A., Pamp, S. J., Lund, O., Hald, T., Woolhouse, M., Koopmans, M. P., Vigre, H., Petersen, T. N., Aarestrup, F. M. Global Monitoring of Antimicrobial Resistance Based on Metagenomics Analyses of Urban Sewage. Nat. Commun. 2019, 10 (1), 1124. https://doi.org/10.1038/s41467-019-08853-3.

(3) García-Aljaro, C., Blanch, A. R., Campos, C., Jofre, J., Lucena, F. Pathogens, Faecal Indicators and Human-Specific Microbial Source-Tracking Markers in Sewage. J. Appl. Microbiol. 2019, 126 (3), 701–717. https://doi.org/10.1111/jam.14112.

(4) Huang, C., Wang, Y., Li, X., Ren, L., Zhao, J., Hu, Y., Zhang, L., Fan, G., Xu, J., Gu, X., Cheng, Z., Yu, T., Xia, J., Wei, Y., Wu, W., Xie, X., Yin, W., Li, H., Liu, M., Xiao, Y., Gao, H., Guo, L., Xie, J., Wang, G., Jiang, R., Gao, Z., Jin, Q., Wang, J., Cao, B. Clinical Features of Patients Infected with 2019 Novel Coronavirus in Wuhan, China. Lancet 2020, 395 (10223), 497–506. https://doi.org/10.1016/S0140-6736(20)30183-5.

(5) Holshue, M. L., DeBolt, C., Lindquist, S., Lofy, K. H., Wiesman, J., Bruce, H., Spitters, C., Ericson, K., Wilkerson, S., Tural, A., Diaz, G., Cohn, A., Fox, L., Patel, A., Gerber, S. I., Kim, L., Tong, S., Lu, X., Lindstrom, S., Pallansch, M. A., Weldon, W. C., Biggs, H. M., Uyeki, T. M., Pillai, S. K. First Case of 2019 Novel Coronavirus in the United States. N. Engl. J. Med. 2020, 382 (10), 929–936. https://doi.org/10.1056/NEJMoa2001191.

(6) Chen, N., Zhou, M., Dong, X., Qu, J., Gong, F., Han, Y., Qiu, Y., Wang, J., Liu, Y., Wei, Y., Xia, J., Yu, T., Zhang, X., Zhang, L. Epidemiological and Clinical Characteristics of 99 Cases of 2019 Novel Coronavirus Pneumonia in Wuhan, China: A Descriptive Study. Lancet 2020, 395 (10223), 507–513. https://doi.org/10.1016/S0140-6736(20)30211-7.

(7) Wu, F., Xiao, A., Zhang, J., Moniz, K., Endo, N., Armas, F., Bonneau, R., Brown, M. A., Bushman, M., Chai, P. R., Duvallet, C., Erickson, W. P., Huang, K. H., Lee, W. L., Matus, M., McElroy, K. A., Nagler, J., Rhode, S. F., Santillana, M., Tucker, J. A., Wuertz, S., Zhao, S., Thompson, J., Alm, E. J. SARS-CoV-2 Titers in Wastewater Foreshadow Dynamics and Clinical Presentation of New COVID-19 Cases. medRxiv 2020, 1–44. https://doi.org/10.1088/1751-8113/44/8/085201.

(8) Bar Or, I., Yaniv, K., Shagan, M., Ozer, E., Erster, O., Mendelson, E., Mannasse, B., Shirazi, R., Kramarsky-Winter, E., Nir, O., Abu-Ali, H., Ronen, Z., Rinott, E., Lewis, Y. E., Friedler, E. F., Paitan, Y., Bitkover, E., Berchenko, Y., Kushmaro, A. Regressing SARS-CoV-2 Sewage Measurements onto COVID-19 Burden in the Population: A Proof-of-Concept for Quantitative Environmental Surveillance. medRxiv 2020, 1–11. https://doi.org/10.1101/2020.04.26.20073569.

(9) Mohan, S. V., Hemalatha, M., Kopperi, H., Ranjith, I., Kumar, A. K. SARS-CoV-2 in Environmental Perspective: Occurrence, Persistence, Surveillance, Inactivation and Challenges. Chem. Eng. J. 2021, 405 (July 2020), 126893. https://doi.org/10.1016/j.cej.2020.126893.

(10) Medema, G., Heijnen, L., Elsinga, G., Italiaander, R., Brouwer, A. Presence of SARS-Coronavirus-2 RNA in Sewage and Correlation with Reported COVID-19 Prevalence in the Early Stage of the Epidemic in The Netherlands. Environ. Sci. Technol. Lett. 2020, 7 (7), 511–516. https://doi.org/10.1021/acs.estlett.0c00357.

(11) Thompson, J. R., Nancharaiah, Y. V., Gu, X., Lee, W. L., Rajal, V. B., Haines, M. B., Girones, R., Ng, L. C., Alm, E. J., Wuertz, S. Making Waves: Wastewater Surveillance of SARS-CoV-2 for Population-Based Health Management. Water Res. 2020, 184, 116181. https://doi.org/10.1016/j.watres.2020.116181.

